# How has the prevalence of smoking cigarettes among HIV-infected patients changed over the last decade? Single-center analysis

**DOI:** 10.1101/2021.02.10.20238949

**Authors:** Pola Tochman, Tomasz Mikuła, Alicja Wiercińska-Drapało

## Abstract

**Introduction:** The WHO report from 2016 estimates the prevalence of smoking as 22% worldwide and 28% in Poland. The prevalence of smoking in HIV-infected persons is 2-3 times higher than in the general population. The study conducted in 2009 in our department among n=116 HIV-infected patients showed 87.1% smoking cigarettes at least 2.5 pack-years (85.0% of males and 93.5% of females). The aim of the present study was to evaluate the change in the prevalence of cigarette smoking among HIV-infected patients over the last decade.

**Methods:** We analyzed data from the survey conducted in n=204 HIV-infected patients hospitalized from November 2018 to November 2019 in our department. The survey included: age; sex, number of cigarettes smoked daily presently and in the past, the number of years as a smoker; the influence of HIV diagnosis on the number of cigarettes smoked.

**Results:** Among all n=204 patients current smokers n=78/204 (38.2%) with n=65/176 (36.9%) males and n=13/28 (46.4%) females. Among all n=115 current smokers and ex-smokers n=29 (25.2%) diagnosing the HIV infection had an impact on their smoking habits: n=26 (89.7%) increased and n=3 - all males (10.3%) decreased the number of smoked cigarettes.

**Conclusion:** The prevalence of smoking cigarettes among HIV-infected individuals in our department has decreased since 2009 but is still much higher than in the general population. The outcome indicates the need to improve smoking education provided by HIV care professionals. Smoking cessation guidance or treatment should become an integral part of HIV care for patients to fully benefit from antiretroviral therapy.

## Introduction

According to the World Health Organization, tobacco use remains one of the major causes of death, killing 8 million people every year [1]. The World Health Organization report from 2016 estimates the prevalence of smoking as 22% worldwide and 28% in Poland (32% of males and 24% of females) [2]. In some populations the prevalence of smoking is still much higher than in general. People living with HIV smoke cigarettes 2-3 times more often than others [3].

In the study conducted in 2009 in our Department of Infectious and Tropical Diseases and Hepatology, Medical University of Warsaw among n=116 HIV-positive patients, 87% had been smokers with at least 2.5 pack-years with 85% of males and 94% of females [4]. Nowadays, due to the advance of antiretroviral treatment, people living with HIV are more likely to die from non-AIDS-related than AIDS-related causes [5]. Tobacco use has a significant contribution to worsening their quality of life and shortening their lifespan [6]. Smoking HIV-positive individuals are more likely than HIV-positive nonsmokers to develop lung, head and neck, and other cancers, heart disease, stroke, chronic obstructive pulmonary disease, bacterial and pneumocystis pneumonia, oral candidiasis, and oral hairy leukoplakia [7].

Among non-AIDS-related death causes, neoplasms are one of the most significant for people living with HIV. According to the research conducted in the United States, seropositive smokers are 6-13 times more likely to die from lung cancer than AIDS-related causes [8]. Studies show that tobacco use results in a poorer response to antiretroviral therapy. Seropositive smokers also experience more difficulty quitting than seronegative people due to higher cotinine blood levels. This association is more significant in females, indicating more serious health consequences and worse results in quit attempts [9].

The aim of our study was to evaluate the prevalence of cigarette smoking among HIV-positive patients in our department compared to results from 2009.

## Methods

We conducted a survey among all HIV-positive patients consulted or hospitalized in the Department of Infectious and Tropical Diseases and Hepatology, Medical University of Warsaw from November 2018 to November 2019. The inclusion criteria were the same for all: adults with confirmed HIV infection, under the care of our department. The exclusion criteria - refusal to participate in the study. The characteristics included: sex; age; present and past smoking habits: the number of cigarettes smoked per day, the number of years as a smoker; the influence of HIV diagnosis on the number of cigarettes smoked. To classify patients as current or ex-smokers we used Centers for Disease Control and Prevention (CDC) National Health Interview Survey (NHIS) criteria: current smoker is an adult who has smoked 100 cigarettes in their lifetime and who currently smokes cigarettes, ex-smoker is an adult who has smoked at least 100 cigarettes in their lifetime but who had quit smoking at the time of the interview, never smoker is an adult who has never smoked, or who has smoked less than 100 cigarettes in their lifetime [10].

The written informed consent was obtained from all participants. This study was approved by the Medical University of Warsaw Bioethics Committee with ID of the approval: AKBE/7/2021.

## Results

We analyzed complete survey data of n=204 (176/28 - males/females) patients with median age 42 (ranges 19–82) years old, mean age 42.9 with standard deviation (SD) 11.6 years. For n=176 males, the median age was 41.5 (ranged 19–82), mean 42.8 with standard deviation 11.9 years. For n=28 females, the median age was 42.5 (ranges 30-65), mean 43.6 with SD – 9.7 years. All baseline data for cigarette smokers are in Table 1.

**Table 1.** Baseline data only for cigarette smokers in 2019**.

|  | **Males** | **Females** |
| --- | --- | --- |
| Total count of cigarette smokers (n/%) | 65/36.9 | 13/46.4 |
| Total count of ex-smokers (n/%) | 32/18.2 | 5/17.9 |
| Number of years as a smoker (median/mean/SD) | 20.0/17.8/9.1 | 15.0/16.2/11.6 |
| Number of cigarettes smoked daily (median/mean/SD) | 20.0/16.0/6.5 | 10.0/13.1/6.0 |

Among all n=115 current smokers and ex-smokers n=29 (25.2%), diagnosing the HIV infection had an impact on their smoking habits: n=26 (89.7%) increased and n=3 - all males (10.3%) decreased the number of smoked cigarettes.

Among all smokers, 25.2% changed their smoking habits after being diagnosed with HIV - 89.7% increased, and 10.3% decreased the number of smoked cigarettes.

Total count of never smokers was n=89 (43.7%) with n=79 (44.9%) males and n=10 (35.7%) females.

After getting our results we compared this data with the results from 2009. We used different definitions for cigarette smokers in 2009 and 2019. In 2009 we defined a cigarette smoker as a person with at least 2.5 pack-years (10 cigarettes per day for 5 years). In 2019 we used different definitions for smokers and ex-smokers according to CDC/NHIS - current smoker is an adult who has smoked 100 cigarettes in their lifetime and who currently smokes cigarettes, ex-smoker is an adult who has smoked at least 100 cigarettes in their lifetime but who had quit smoking at the time of interview. The definition of never smoker was similar in both surveys: in 2009 it was the patient who has never smoked before and in 2019 we used the definition for never smoker as an adult who has never smoked, or who has smoked less than 100 cigarettes in their lifetime.

The percentage for all smokers, males and females are presented in Figure 1 (see Figure 1).

**Figure 1.**
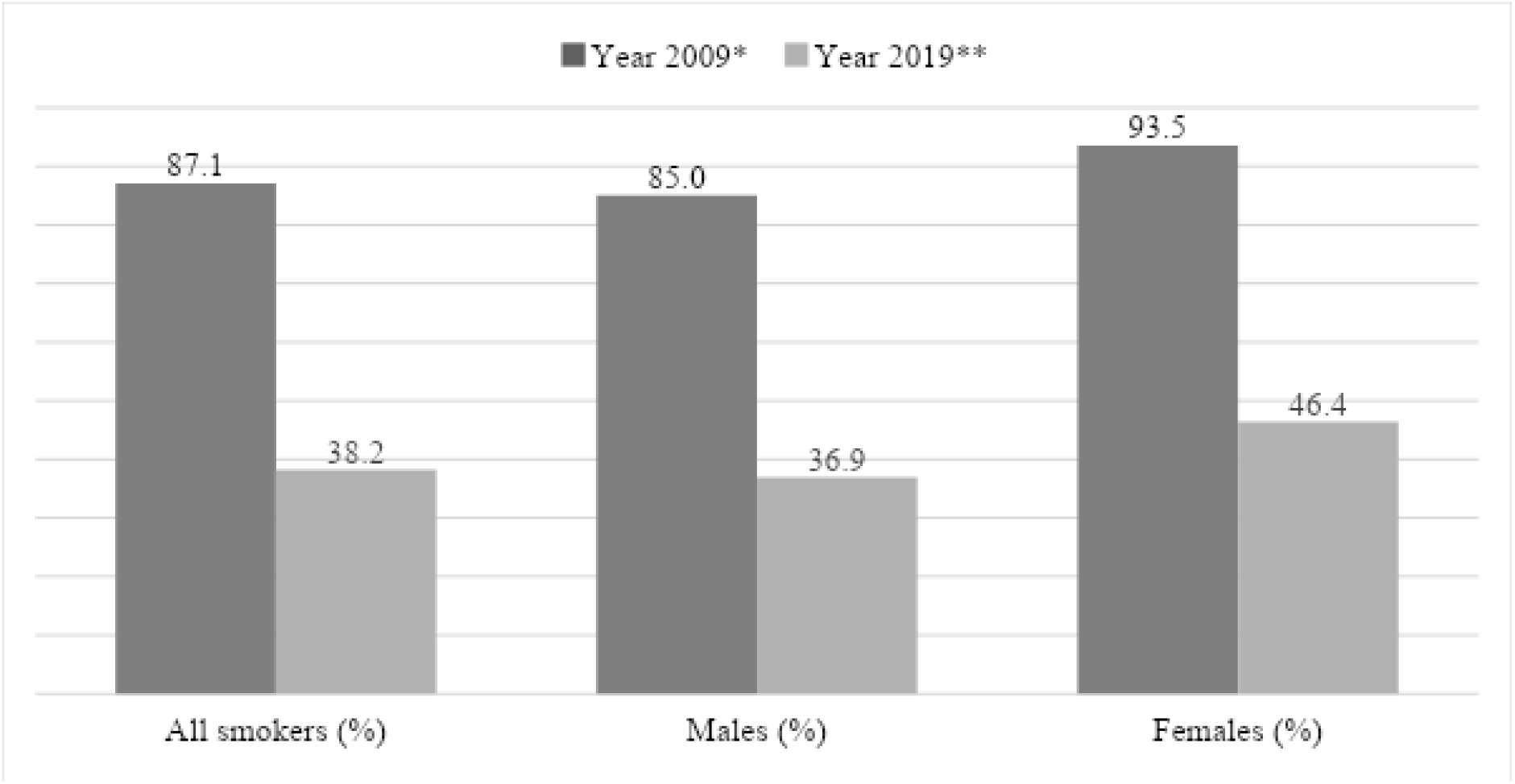
The percentage (%) of cigarette **smokers** among HIV-infected patients in analyzed years 2009* and 2019**. *In 2009 we used the definition: cigarette smoker was a person with at least 2.5 pack-years (10 cigarettes per day for 5 years). **In 2019 we used the definition: cigarette smoker was a person who has smoked 100 cigarettes in their lifetime and who currently smokes cigarettes.

The percentage for all never-smokers, males and females are presented in Figure 2 (see Figure 2).

**Figure 2.**
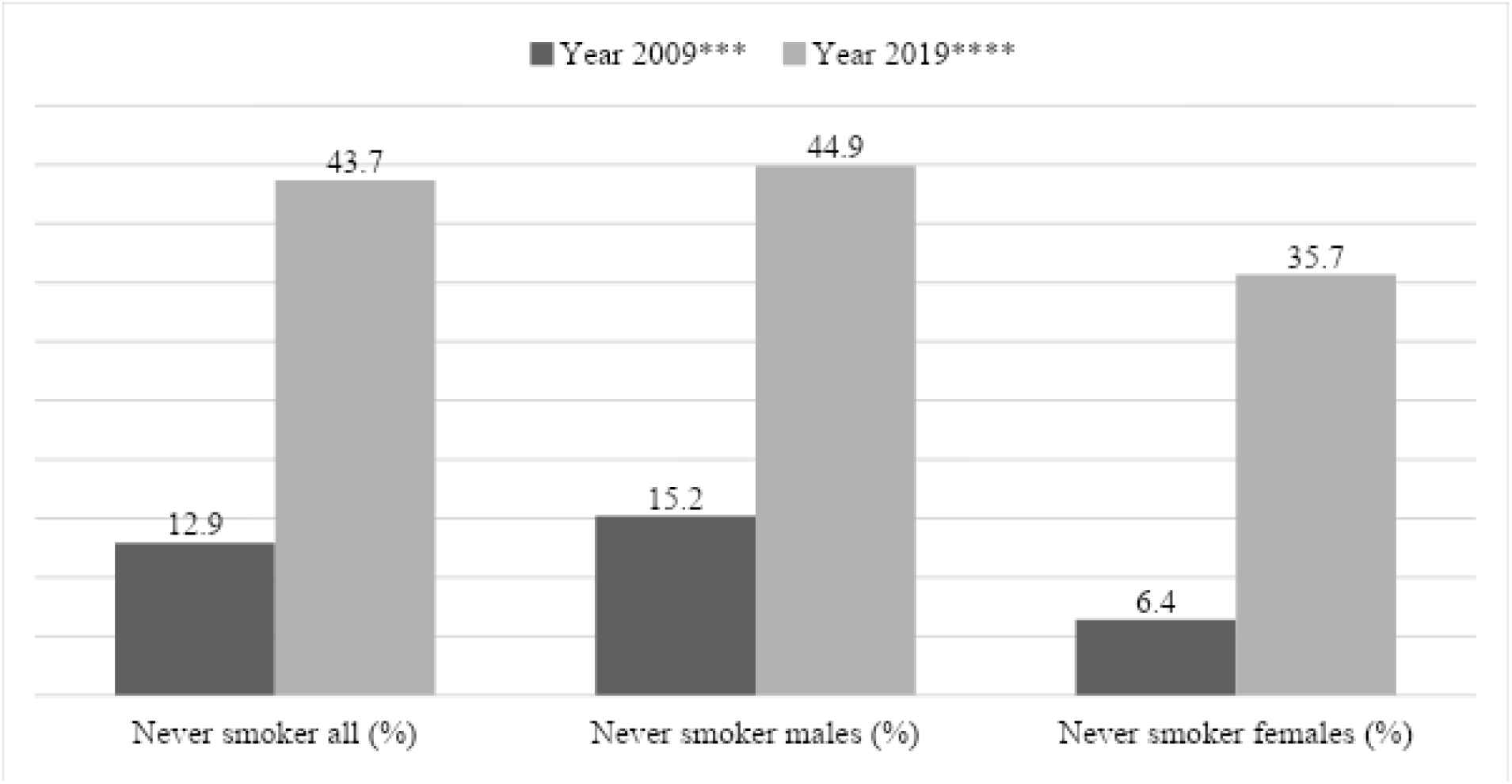
The percentage (%) of cigarette **never-smokers** among HIV-infected patients in analyzed years 2009*** and 2019****. ***In 2009 we used the definition: never-smoker has never smoked before. ****In 2019 we used the definition: never-smokers as an adult who has never smoked, or who has smoked less than 100 cigarettes in their lifetime.

## Discussion

In Poland, in the period from 1985 to 30 September 2019, there were 25 020 cases of confirmed HIV infection, with 3 741 patients diagnosed with AIDS among whom 1 424 died but still, there are not enough studies assessing the scale of tobacco use in this population [11].

In the study conducted in 2015 in the United States among 419 945 patients, 42.4% were current smokers, 20.3% were ex-smokers, and 37.3% had never smoked, while in the general population in this country 20.6% of adults smoked. HIV-positive individuals who smoke tobacco were also less likely to quit than HIV-negative smokers [12].

A study by Hellenberg et al. conducted in Denmark shows that people living with HIV underestimate the harmful effects of tobacco use. Self-perceived life expectancy among smokers was 3.65 years lower than among non-smokers, while studies show that smoking HIV-positive individuals have 12.3 years of life lost compared to HIV-positive non-smokers. Participants estimated life lost attributable to the HIV status as least as strong as due to current smoking, however, researchers found tobacco use causing more than twice the number of years lost than HIV infection alone. Moreover, the other Danish study by Ronit et al. confirmed the male sex was associated with a lower self-perceived life expectancy [6,13].

Comparing to data from other high-income countries, the prevalence of smoking among those patients in Poland seems to follow the trend: people living with HIV are more than twice as likely to smoke than others.

Over the last decade, we can observe the significant reduction of smoking cigarettes among HIV-infected patients in our department from 87.1% in 2009 to 38.2% in 2019. We should note the difference in the methods used (the definition of the smoker), which suggests that in 2009 the percentage of smokers was even higher than presented. The percentage of smokers decreased in males from 85.0% to 36.9% and in females from 93.5% to 46.4%. It may be related to the suggestion that HIV-positive females experience more difficulty quitting [9]. However, in the study conducted in China among n=2973 people living with HIV where 28.9% of participants reduced, 19.0% quit and only 8.2% increased smoking after being tested HIV-positive, female sex was associated with quitting [14].

In our data from 2019, we observed 43.7% never smokers with 44.9% among males and 35.7% among females. Of all smokers, 25.2% changed their smoking habits after being diagnosed with HIV: 89.7% increased and 10.3% decreased the number of smoked cigarettes. Only 18.2% of males and 17.9% of females decided to stop smoking during HIV infection.

Our results show that diagnosing HIV infection plays a role in changing smoking habits. The fact that a small group of patients decided to reduce or quit cigarettes after being diagnosed indicates the absence of sufficient, effective smoking education carried out by HIV care providers.

The results of another polish study by Janik-Koncewicz et al. show a strong need to support cessation treatment, although research conducted by the Health Promotion Foundation in 2016 indicates the insufficient level of knowledge in this area among polish HIV care providers. Doctors and nurses working with people living with HIV were asked about tobacco-related diseases, psychological support during cessation treatment, available cessation medicines. Researchers also evaluated how often doctors talk with their patients about smoking. Results show that HIV care professionals in Poland lack the essential knowledge needed to provide effective cessation treatment and often ignore this topic during medical visits. Respondents highlighted that the guidelines of the Polish AIDS Society should be expanded in the matter of smoking treatment [15]. Providing cessation treatment for seropositive smokers should be an integral part of HIV care and one of its priorities. Because of smoking cigarettes, HIV-infected patients are not able to fully benefit from antiretroviral therapy, which nowadays increases life expectancy in people living with HIV to even equal to those in the general population [16].

## Conclusion

The prevalence of smoking cigarettes among HIV-infected individuals in our department has decreased since 2009 but is still much higher than in the general population. It is necessary to underline the need to improve smoking education and guidance provided by HIV care professionals. Talking about smoking cigarettes cessation or treatment should become an integral part of HIV care for patients to fully benefit from antiretroviral therapy.

## Data Availability

The authors confirm that the data supporting the findings of this study are available within the article.

## Acknowledgements

Study concept, survey design, data collection and analysis: P.T., T.M.

Writing: P.T.

Editing and review: all authors.

## Data sharing statement

All data relevant to the study are included in the article.

